# Socio-demographic determinants of COVID-19 vaccine uptake in Ontario: Exploring differences across the Health Region model

**DOI:** 10.1101/2023.08.04.23293662

**Authors:** Ariel Mundo Ortiz, Bouchra Nasri

**Affiliations:** Centre de Recherches Mathématiques, University of Montreal, Montréal, Canada; Department of Social and Preventive Medicine, École de Santé Publique, University of Montreal, Montréal, Canada; Centre de recherche en santé publique, University of Montréal, Montréal, Canada

**Keywords:** COVID-19, vaccination, survey, socio-economic factors, visible minorities, race and ethnicity

## Abstract

The COVID-19 pandemic continues to be a worldwide public health concern. Although vaccines against this disease were rapidly developed, vaccination uptake has not been equal across all the segments of the population. In particular, it has been shown that there have been differences in vaccine uptake across different segments of the population. However, there are also differences in vaccination across geographical areas, which might be important to consider in the development of future public health vaccination policies. In this study, we examined the relationship between vaccination status (having received the first dose of a COVID-19 vaccine), and different socio-economic and geographical factors. Our results show that between October of 2021 and January of 2022, individuals from underrepresented communities were three times less likely to be vaccinated than White/Caucasian individuals across the province of Ontario in Canada, and that in some cases, within these groups, individuals in low-income brackets had significantly higher odds of vaccination when compared to their peers in high income brackets. Finally, we identified significantly lower odds of vaccination in the Central, East and West Health Regions of Ontario within certain underrepresented groups. This study shows that there is an ongoing need to better understand and address differences in vaccination uptake across diverse segments of the population of Ontario that the pandemic has largely impacted.

## Introduction

As of May of 2023, there have been 765 million confirmed cases of COVID-19 around the world, including 6.8 million deaths^1^. Although this disease is no longer categorized as a global health emergency by the World Health Organization (WHO)^2^, there is ongoing concern due to continued transmission, the possibility of surges in cases and deaths due to new virus variants^3^, and ongoing issues in health systems around the world that could be exploited by a novel virus or another public health emergency in the future^4^.

In particular, the pandemic showed multiple challenges with regard to vaccination. The rapid development of vaccines against COVID-19 initially brought the hope of a rapid end to the pandemic as vaccination campaigns in certain parts of the world started as early as December of 2020^5–8^. Although it has been estimated that so far COVID-19 vaccines have been able to prevent millions of deaths worldwide^9^, their implementation has faced multiple challenges across the world. These challenges included the rapid emergence of new virus variants^10^, the waning of vaccine protection^11^, inequality in vaccine access between high-income and low-income countries^12,13^, vaccine hesitancy^14^, and differences in vaccine uptake across the population^15,16^. More specifically, it is well established that lower vaccination uptake has been observed in individuals within certain underrepresented groups (e.g., Black, Asian, or Indigenous) as well as individuals with socio-economic disadvantages^15–21^.

Reasons given for this inequality in vaccination uptake have included medical mistrust due to systemic medical racism^17,22^, mistrust in vaccines^15^, and the existence of health misinformation and disinformation^22–24^. However, it is important to consider that vaccination uptake can also vary geographically across the administrative or political subdivisions within a country. For example, differences in COVID-19 vaccination rates have been associated with differences in attitudes towards vaccination in administrative areas in New York and Chicago^25^, scarcity of vaccination facilities in areas where underrepresented groups reside in Toronto when the provincial vaccination program started^7^, as well as an absence of prioritization of areas inhabited by vulnerable groups in the southeastern region of the US^26^. Other studies have also shown heterogeneity in vaccine uptake within counties in the US^27–30^, indicating that accounting for geographical differences in vaccination can help predict patterns of booster uptake in Scottish communities^31^. Therefore, the study of vaccination uptake across administrative of political areas can be useful to highlight differences that governments or public health agencies might need to address. However, in the case of Canada, there is a limited amount of studies that have analyzed geographical differences in vaccination. Existing studies in this area have focused on differences within certain cities, such as Toronto^32^, or Montréal^33^), or have explored differences between provinces-at-large^19^, but to our knowledge, there are no studies that analyze differences in vaccination uptake at the intra-provincial level. Such need is specially important in the context of Canada’s pandemic response goals, which have been to minimize serious illness and deaths while minimizing societal disruption^34^. Analyzing differences in vaccination uptake within the provinces can aid to identify inequalities that might exist and that need to be addressed before the advent of another pandemic.

This need is especially important in the case of Ontario, the most populated province of Canada, and which as has a complex healthcare system. Between 2007 and 2019, Ontario managed healthcare access to its inhabitants using 14 intra-provincial divisions called the Local Health Integration Networks (LHINs), which aimed to provide an integrated health system. However, this approach was complex, bureaucratic, resulting in excessive expenditures, disparities in mortality rates, the deterioration of certain performance indicators (such as wait times and hospital readmissions), fragmented electronic health systems, and inequities in health services access^35–39^. With the intent of better organizing and delivering care, in late 2019 the provincial government eliminated the LHINs and incorporated the areas covered by them into six larger Health Regions (North East, North West, Central, Toronto, West, and East), which are managed by a new government agency, Ontario Health (OH)^37^.

On the other hand, public health in Ontario is administered by Public Health Ontario (PHO), a government agency established in late 2007 and that is currently composed of 34 Public Health Units (PHUs) that cover the entire geography of the province. Although PHUs were commissioned with leading the distribution of the COVID-19 vaccine within their respective areas^40^, the vaccine rollout occurred with significant interaction between PHO and OH; in many instances both agencies had to work together to organize vaccination clinics, with personnel associated with OH also being actively involved when demand exceeded the capacity of PHO personnel, especially in rural areas^41^. Indeed, based on the experience of COVID-19, there are ongoing discussions on how OH and PHO will interact in the future, and the challenges that this will entail for the healthcare system of the province^42^.

Therefore, considering the relatively recent adoption of the Health Region model and its alignment with the onset of the COVID-19 pandemic, there is an ongoing need to analyze the existence of geographical disparities in vaccination uptake within the Health Regions and identify the socio-demographic groups that might be affected, as this can serve as an indirect assessment of the state of the implementation of the new model while helping identify ongoing challenges that decision-makers might need to address to ensure the long-term success of this model and its interaction with PHO, which is specially important considering that previous research has highlighted disparities in the level of activity of each Health Region^43^.

Therefore, in this study we wanted to understand if there were differences in COVID-19 vaccination rates among the Health Regions between October 2021 and January of 2022. To contextualize these differences, we included socio-economic factors in our analysis aiming to identify which demographic groups were particularly impacted, in order to provide an assessment of the current state of healthcare access in Ontario.

## Methods

### Data and Methods

We used data from the *Survey of COVID-19 related Behaviours and Attitudes*, a repeated cross-sectional survey focused on the Canadian province of Ontario that was commissioned by the Fields Institute for Research in Mathematical Sciences and the Mathematical Modelling of COVID-19 Task Force under ethical guidance from the University of Toronto (under Research Ethics Board approval #40999), and which ran between September 30th, 2021 and January 17th, 2022.

Briefly, the survey was deployed using Random Domain Intercept Technology (RDIT), a methodology for internet surveys developed by the company commissioned to run the survey (RIWI Corp., Toronto, Canada), and that has been used in the area of public health research to examine trends in vaccination rates^44^, ratings of care quality^45^, and perceptions of vaccine efficacy^46^. In the case of the survey used for this study, internet users whose device metadata indicated their presence in the province of Ontario had a random chance of being redirected to the survey after they had clicked on a registered but commercially inactive web link, or after they had typed in a web address for a site that was dormant but that was temporarily managed by RIWI. Users then decided whether to anonymously participate in the survey, and those that participated were able to exit the survey at any time. After the survey closed, regardless if it was complete or incomplete, access was denied to any further users with the same internet protocol address (IP), effectively allowing each user only one opportunity to participate in the survey. Users who indicated they were under the age of 16 were exited from the survey without creating a record. Finally, the personal identifier information from each user that participated in the survey was automatically scrubbed and replaced by a unique ID.

Survey users entered their socio-economic information (age, income, and racial/ethnic group), and were asked information on vaccination status by using the question “Have you received the first dose of the COVID vaccine?”, with possible answers “yes” and “no” (Table 1). Of notice, the racial/ethnic and socio-economic categories from the survey did not match exactly the categories from the 2016 Census (e.g., the categories “East Asian/Pacific Islander” as well as the income bracket “over CAD 110,000” do not exist in the Census data). Therefore, we used a combination of sources to re-group certain categories and socio-economic strata in order to obtain estimates from the Census that could be used to correct the data. Further details can be found in the Appendix.

**Table 1:** Descriptive Statistics of the Fields COVID-19 Survey (by Vaccination Status)

| Variable | no, N = 1,547<sup>1</sup> | yes, N = 4,689<sup>1</sup> |
| --- | --- | --- |
| Income (CAD) |  |  |
| 60000 and above | 542 (21%) | 1,996 (79%) |
| 25000-59999 | 347 (25%) | 1,046 (75%) |
| under 25000 | 658 (29%) | 1,647 (71%) |
| Age Group |  |  |
| 16-34 | 645 (23%) | 2,117 (77%) |
| 35-54 | 411 (24%) | 1,305 (76%) |
| 55 and over | 491 (28%) | 1,267 (72%) |
| Health Region |  |  |
| Toronto | 593 (26%) | 1,709 (74%) |
| Central | 372 (26%) | 1,083 (74%) |
| East | 236 (23%) | 783 (77%) |
| West | 346 (24%) | 1,114 (76%) |
| Month |  |  |
| October 2021 | 469 (27%) | 1,263 (73%) |
| November 2021 | 376 (28%) | 980 (72%) |
| December 2021 | 181 (24%) | 565 (76%) |
| January 2022 | 521 (22%) | 1,881 (78%) |
| Race |  |  |
| White/Caucasian | 354 (16%) | 1,871 (84%) |
| Arab/Middle Eastern | 111 (34%) | 220 (66%) |
| Black | 159 (34%) | 303 (66%) |
| East Asian/Pacific Islander | 94 (19%) | 404 (81%) |
| Indigenous | 112 (37%) | 194 (63%) |
| Latin American | 99 (34%) | 195 (66%) |
| Mixed | 177 (30%) | 411 (70%) |
| Other<sup>2</sup> | 315 (34%) | 606 (66%) |
| South Asian | 126 (21%) | 485 (79%) |

<sup>1</sup>n (%)
<sup>2</sup>This category included Southeast Asian, Filipino, and West Asian individuals, and minorities not identified elsewhere according to the 2016 Census.

Additionally, the survey automatically collected the geographical location of the respondent (using the nearest municipality, as shown in Figure 1), and the date of access to the survey. The original dataset contained 39,029 observations. However, the number of complete observations was much lower than the total number of observations due to the survey design, which allowed respondents to exit at any time and deployed the questions randomly. We selected the observations with complete answers (6,343 observations, or 16.25% of the total) for our analysis. It should be noted that this response rate is similar to response rates observed in previous studies that have used the same type of survey instrument, with response rates between 15% and 22%^45,46^. Next, we matched the city of each observation with its corresponding LHIN and Health Region, and removed observations from areas with low representation (254 observations corresponding to the North West and North East Health Regions). After all the preliminary analyses, the total number of observations used for analysis was 6,236 and included the East, Central, Toronto, and West Health Regions covering between October 1st, 2021 and January 17, 2022. The original dataset, clean dataset, and details on the data cleaning process, and data preparation are described in detail in the GitHub repository for this paper.

**Figure 1:**
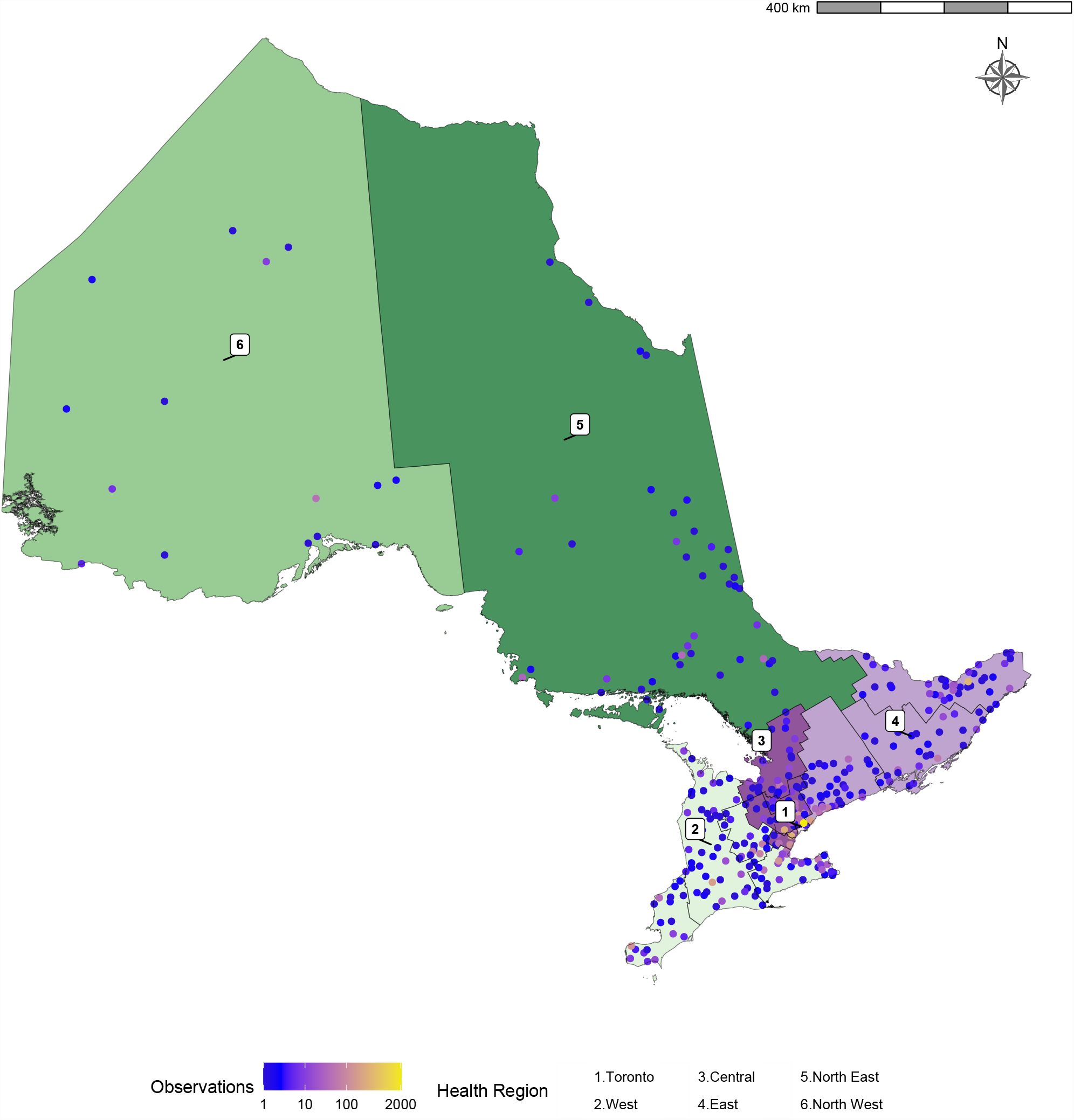
Geographic representation of the data collected by the *Survey of COVID-19 related Behaviours and Attitudes*, collected by the Fields Institute in Ontario. Municipalities from where survey participants provided answers appear as points, color indicates number of observation obtained from each city. The Health six Regions are color-coded and labelled sequentially. Internal boundaries within certain Health Regions indicate areas previously covered by the Local Integrated Health Networks (LHINs).

To analyze the survey data, we used a logistic regression model to examine the impact of the Health Regions in the uptake of the first dose of the vaccine, using as the variable of interest the question “Have you received the first dose of the COVID vaccine?” (with possible answers “yes” or “no”), while considering as independent variables the socio-economic factors of the participants, months covered by the survey (Table 1), and the Health Regions of OH. Furthermore, we included certain interactions (Race and Health Region and Race and income) in our model as previous studies have shown that socio-economic factors and their interactions are significant predictors of intent of vaccination and vaccination status^47–49^.

Because we identified differences between the percentage of participation within each of the socio-demographic variables collected by the survey that were considered in our analysis and the percentage within these factors in the 2016 Census (such as participation by age and racial/ethnic groups), we used an iterative proportional fitting procedure (*raking*)^50^ to correct the percentages within each socio-demographic variable using population totals obtained from the Census and OH; later, we fitted the regression model to the uncorrected and corrected data to determine if there were any differences in the obtanied estimates. Details regarding the correction can be found in the Appendix. All analyses were conducted in R 4.2.2 using the packages survey^51^,tidyverse^52^, quarto^53^, modelsummary^54^, and gtsummary^55^.

## Results

### Sample Characteristics

Table 1 shows the characteristics of the data from the Fields COVID-19 survey used for analysis. The sample contained 6,236 observations, from which 24.8% (1,547) corresponded to individuals that reported not having initiated a COVID-19 vaccine primary series (in other words, not having received the first dose of the vaccine). The rate for the first dose of the vaccine ranged between 71-79% across all household income brackets, age groups, Health Regions, and the months considered in the survey. However, the highest rate for the uptake of the first dose of the vaccine were reported by individuals in the highest income bracket (79%), those between 16 and 34 years of age (77%), individuals that lived in the East Health Region (77%), and during January of 2022 (78%). Between racial/ethnic groups, White/Caucasian individuals reported the highest uptake of the first dose of the vaccine (84%), against values that ranged between 63 and 66% in the case of Arab/Middle Eastern, Black, Indigenous, Latin American individuals, and those that belonged to the “other” racial group category (which included Southeast Asian, Filipino, West Asian, and minorities not identified elsewhere percentages).

### Multivariate Regression

Figure 2 presents the estimates (as odd ratios) from the logistic regression models for vaccination status using the socio-demographic factors collected by the survey, and their interactions for the corrected and uncorrected data. Generally speaking, lower odds of vaccination were identified in both cases in individuals characterized by a low household income, or that identified as part of underrepresented groups. However, the magnitude of the estimates differed between the uncorrected and corrected models and more importantly, there were differences in the statistical significance of certain estimates before and after the correction. Specifically, the uncorrected model showed significant differences in vaccination odds between the age groups considered, the East Health Region, Latin American individuals with a household income under CAD 25,000, and Indigenous individuals living in the Central Health Region (Figure 2,B) but these were deemed non statistically significant after the correction.

**Figure 2:**
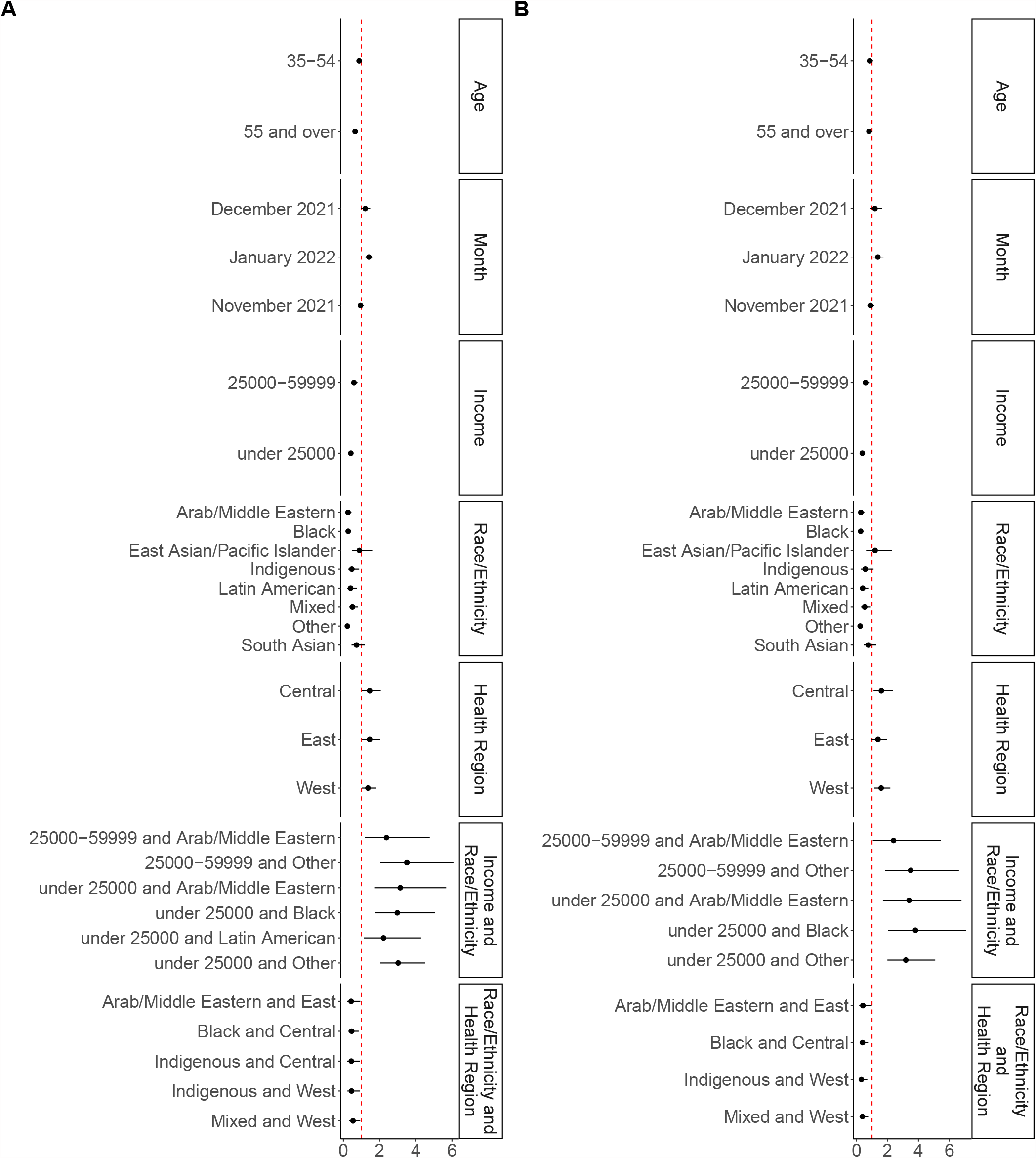
**A**. Coefficient estimates and confidence intervals for the uncorrected model. **B**. Coefficient estimates and confidence intervals for the corrected model. Only statistically significant interaction terms are shown in both cases. Full interaction terms can be found in Supplementary Figures A-3 and A-4.

However, significantly lower odds of vaccination were identified in the corrected model for those with a household income under CAD 25,000 (OR=0.37, CI=[0.27,0.51]) and those with an income between CAD 25,000 and 59,999 (OR=0.58, CI=[0.42,0.81]). Additionally, individuals who identified as Arab/Middle Eastern, Black, Latin American, of mixed background, or that belonged to other racial groups (a category that included Southeast Asian, Filipino, West Asian, and minorities not identified elsewhere), had significantly lower odds of vaccination than those in the White/Caucasian group (ORs and CIs=0.28 [0.16,0.51], 0.27 [0.16,0.45], 0.40 [0.21,0.76], 0.53 [0.30,0.92], 0.23 [0.15,0.36]). Additionally, individuals that reported living in the Central and West Health Regions had higher odds of vaccination than those in the Health Region of Toronto (ORs and CIs=1.61 [1.10,2.34], and 1.59 [1.16,2.19], respectively).

Interestingly, individuals in underrepresented groups with a household income below CAD 25,000 had higher odds of vaccination (when compared to those with a household income above CAD 60,000). This held true in the case of Arab/Middle Eastern individuals (OR=34, CI=[1.70,6.79]), Black individuals (OR=3.81, CI=[2.05, 7.09]), and those in other racial or ethnic groups (OR=3.19, CI=[2.00,5.09]). Additionally, individuals with an income between CAD 25,000 and 59,999 in the Arab/Middle Eastern and other racial or ethnic groups also had higher odds of vaccination than their high-income peers (ORs and CIs=6.96 [2.67,18.16], and 3.5 [1.85,6.62]).

Finally, the place of habitation affected the odds of vaccination for certain underrepresented groups, as significantly lower odds of vaccination were identified for the interaction between Health Region and race in the case of Black individuals in the Central Health Region (OR=0.39, CI=[0.2,0.75]), Arab/Middle Eastern individuals in the East Health Region (OR=0.41 [0.17, 0.98]), and in the Indigenous and mixed groups in the West Health Region (ORs and CIs=[0.31 [0.14, 0.7] and 0.38 [0.19, 0.76], respectively).

## Discussion

In this study, we wanted to understand if there were differences in COVID-19 vaccination rates among the Health Regions between October 2021 and January of 2022 while identifying the socio-demographic determinants (and their interactions) that could help predict these differences. Using the Health Regions as the base of our analysis was advantageous, as these newly implemented administrative areas match closely the geographical boundaries of the Health Regions that have been historically used to group Public Health Units in the province^56,57^. Thus, a Health Region-based analysis could provide information on vaccination uptake that could be useful to understand patterns within the realm of PHO and OH. Moreover, such analysis could provide an overall assessment of intra-provincial disparities that might need to be addressed moving forward by decision-makers in order to ensure that in the event of a future public health emergency or pandemic, OH and PHO will able to work collaboratively in an efficient way having learned from the COVID-19 experience^41^, thus ensuring that the population of the province benefits in the long term from a robust health system.

Our results show that indeed, there were differences in the uptake of the first dose of the vaccine across Ontario in certain socio-demographic groups. Specifically, those who identified as Arab/Middle Eastern, Black, Latin American, having mixed racial or ethnic background, or that belonged to other groups not explicitly included in the survey (Southeast Asian, Filipino, West Asian, and minority groups not identified elsewhere) had vaccination odds that were between a third and a half of that of individuals that identified as White or Caucasian (Figure 2). These results are consistent with previous studies that have shown lower vaccination rates in individuals with the same socio-demographic characteristics^19–21,58^.

Lower vaccine uptake in the socio-demographic groups indicated above may be influenced in part, by vaccine hesitancy and refusal, which have been associated in underrepresented Canadian individuals with concerns on vaccine safety, effectiveness, and experiences of racial discrimination in health settings^49,59–61^. However, it has been shown that structural barriers also play an important role in vaccination uptake. In the case of underrepresented individuals, such barriers include complex scheduling systems, language barriers, lack of adequate public transportation, and lack of accessible vaccination sites^62^. In this regard, it is interesting to note that vaccination venues were scarce in low socio-economic areas that had the highest burden of COVID-19 in Toronto and other regions of Ontario around the time covered by the survey^7,63^, and that pharmacies in the Peel region (an area identified as a “hotspot” with high numbers of essential workers and multigenerational households) could not keep up with vaccine demand^64^. This suggests that the observed differences are associated with disparities in vaccine access that were present during the period covered by the survey.

Interestingly, whereas overall self-reported vaccination rates were found to be statistically significantly lower in various underrepresented groups when compared to White/Caucasian individuals, the change in odds of vaccination within certain racial groups and income strata was actually positive, in contrast to the White/Caucasian group, where vaccination odds decreased in income brackets below CAD 60,000 (Supplementary Figure A-5). Specifically, individuals in low income brackets that belonged to Arab/Middle Eastern, Black, or other minority groups had higher odds of vaccination that their peers with an income above 60,000 CAD.

This result likely reflects in part the fact that individuals in underrepresented groups tend to perform occupations that have been deemed as “essential” in the context of the pandemic^65,66^, which include workers in the areas of grocery stores, gas stations, warehouses, distribution, and manufacturing, all being occupations for which an income within the significant brackets identified in the analysis is to be expected. From one side, individuals in essential occupations in the province experienced higher rates of morbidity and mortality during the first year of the pandemic^67^, but later on, they had priority for COVID-19 vaccination^68^. Additionally, it is known that vaccination uptake in these individuals was encouraged by vaccination staff in certain parts of the province^64^. These facts, combined with evidence of increased trends in vaccination in this group elsewhere^69^, suggest that in Ontario, the type of occupation of individuals in underrepresented groups (which might have also affected their decision to get a vaccine based on their knowledge of increased risk), played an important role in the higher the odds of vaccination observed in these individuals.

However, the results also indicate that the place of habitation affected the odds of vaccination for certain under-represented groups (interaction term of Health Region and Race, Figure 2,B). Specifically, this held true in the case of individuals identifying as Indigenous or with mixed racial background in the West Health Region, Black individuals in the Central Health Region, and Arab/Middle Eastern individuals in the East Health Region Figure 2. For these individuals, vaccination odds were lower when compared to the Toronto Health Region (Supplementary Figure A-6). We indicate next some contributing factors that might help provide context to these results.

First, in this case it is useful to analyze the data considering the LHINs in each Health Region, because most studies in the literature focused on Ontario use the LHINs as the base of their analyses. The West Health Region covers the area previously occupied by the Hamilton Niagara Haldimand Brant, South West, and Waterloo Wellington LHINs, whereas the East Health Region covers the area of the former Champlain and Central East LHINs. Previous research has identified health disparities in these (mostly rural) regions, such as unequal distribution of primary care providers, increased mortality, and low pharmacist availability^70–72^.

Furthermore, there is an ongoing challenge for the health system of the province with regard to personalized healthcare for marginalized individuals. Indeed, previous studies have pointed out mistrust in the traditional healthcare system (due to systemic racism or oppression) as a rationale for lower vaccination rates in the case of Indigenous and Black individuals in Canada^73,74^. Other studies have also shown a need for increased culturally responsive care for other underrepresented groups such as Black women and and other immigrant groups in Canada and Ontario^75,76^. Considering that for example, the West Health Region has only two Aboriginal Health Access Centres (community-led primary healthcare organizations focused on First Nations, Métis, and Inuit communities) to provide care to an estimated population of 100,000 Indigenous individuals living in the area^77^, it is possible that the limited personalized healthcare for underrepresented groups in certain parts of the province impacted vaccination uptake as well, and highlights the need of investments in the Health Regions focused on resources, infrastructure, and specially personnel that can deliver personalized care to marginalized communities, as it has been shown that such efforts have improved trust in vaccination in underrepresented groups elsewhere^78^.

There are some limitations to the present study. First, the data collection design, which allowed respondents to withdraw from the survey at any point, and that deployed the questions in a random manner resulted in an elevated number of missing observations without a definite pattern and complicated the implementation of sensitivity analyses. Therefore, we focused on entries that had complete answers, and corrected the data using population-wide information from the Census. However, more granular corrections would be needed to obtain more accurate estimates: For example, our analysis identified higher odds of vaccination in the Central and West Health Regions, but in this case these differences are likely to be driven by the proportion of White/Caucasian individuals, who had higher vaccination rates than other racial groups. Correcting for each racial/ethnic group in each Health Region would provide a more accurate estimation of region-wide vaccination rates but unfortunately, presently this correction cannot be implemented as such stratification has not been implemented in the open data that can be obtained from the Census.

Additionally, our analysis did not consider the North West and North East Health Regions, due to the low number of entries from these areas in the survey (Figure 1). Low representation is expected as these regions as they only account for 5% of the total population of Ontario. However, these areas have the highest proportion of Indigenous inhabitants^77^. In the context of personalized care, there is a need to collect data that focuses on these Health Regions where additional health disparities might be present and possibly understudied.

The results in this study are based on self-reported data, where bias might be present. However, in the context of COVID-19, it has been shown that good agreement exists between self-reported and documented vaccination status^79^, we believe that our data was able to provide a valid sample of vaccination uptake in the province. This is supported by the statistically significant higher vaccination odds that were identified for January of 2022 in the model, which are consistent with province-wide trends reported by Public Health Ontario (which show a 4% increase between early December and January, in contrast to a 2.5% increase between October and November^80^); however, the short time window constitutes essentially a “snapshot” view of the evolution of the disease, and additional data would be needed to obtain estimates per racial/ethnic group over time across all Health Regions that can help inform the existence of other health disparities.

Nonetheless, the results presented here can serve as a starting point to motivate the collection of robust longitudinal data that can be used to quantify geographical and temporal differences within vulnerable segments of the population, and that can be used to inform the development of adequate public health policies within the province of Ontario or across other provinces in Canada that aim to minimize disparities in health access.

## Conclusion

The evidence collected in this study shows differences in the uptake of the first dose of the COVID-19 vaccine in Ontario between October 2021 and January 2022 in underrepresented groups, which had significantly lower odds of vaccination when compared to White/Caucasian individuals. However, although overall vaccination uptake was lower in underrepresented individuals in the province as a whole, the odds of vaccination differed within certain income levels and Health Regions in these groups. Specifically, those that reported a low household income had higher vaccination odds when compared to individuals in the same racial/ethnic group in a higher income bracket. These results highlight the complex landscape of the province, where varying degrees of rurality exist in conjunction with a socio-demographic makeup that is unique to each of the Health Regions.

Personalized care is an area that could be further developed to improve vaccination uptake in underrepresented individuals in the future. As we have shown in the discussion, there are currently a limited number of centers focused on community-led healthcare in the province. Our results showed the existence of differences in vaccination uptake between certain Health Regions and therefore, improving personalized care for underrepresented individuals can serve as a point to improve trust and facilitate vaccine access in these marginalized communities.

Overall, there are ongoing challenges for healthcare access across different segments of the population of Ontario. Future studies can be focused at more in-depth analyses of vaccination uptake within the different socio-demographic groups in each Health Region, in order to provide decision-makers with information that can serve to carefully consider and address how to improve vaccine access to marginalized communities in Ontario in the event of a future public health emergency.

## Data Availability

The survey data used in this study was collected by the Fields Institute for Research in Mathematical Sciences. Those interested in accessing the survey data used in this study should contact directly the Fields Institute for Research in Mathematical Sciences. Data used for corrections, and the mapping of observations to their corresponding Health Regions can be found in the GitHub repository for this work.

https://github.com/aimundo/COVID-19_vaccination/

## Acknowledgments

This work was supported by the Fonds de recherche du Québec Scholar Program (J1 in Artificial Intelligence and Digital Health, BN), the Natural Sciences and Engineering Research Council of Canada through the Discovery Grant Program (RGPID-560523-2020, BN), the Mathematics for Public Health (MfPH) Emerging Infectious Diseases Modelling Initiative (BN, AM), and the OMNI Emerging Infectious Disease Modelling Initiative (BN).

The authors thank Dr. Sarah Wilson, Medical Epidemiologist with Public Health Ontario, for her valuable comments and feedback.

