## Supplementary File: Appendix for "Socio-demographic determinants of COVID-19 vaccine uptake in Ontario: Exploring differences across the Health Region model"

This document provides details on assessing the socio-economic information from the clean survey dataset, and how it compared to the Census Data for ontario. At the end, it also provides information about the corrections implemented into the data. The variables examined were: race/ethnicity, age groups, income, and population by Health Region. Population-wide totals were obtained from the 2016, available in the Statistics Canada Website.

#### Race and Ethnicity

First, we explored how the race and ethnicity information from the dataset and how it compared to the data from the Census for Ontario. The following chunk creates a summary table for the Race variable.

Table A-1: Ethnic information from the clean dataset

| Race | observations | percentage |
|---|---|---|
| White/Caucasian | 2225 | 35.7% |
| Arab/Middle Eastern | 331 | 5.3% |
| Black | 462 | 7.4% |
| East Asian/Pacific Islander | 498 | 8.0% |
| Indigenous | 306 | 4.9% |
| Latin American | 294 | 4.7% |
| Mixed | 588 | 9.4% |
| Other | 921 | 14.8% |
| South Asian | 611 | 9.8% |

It is important to mention that the categories for race/ethnicity provided in the survey did not match the categories used in the Census. Therefore, we used a combination of sources to obtain estimates of racial/ethnic distribution that matched the categories from the survey.The data sources were:

- Fact sheet from the Provice of Ontario for Visible Minorities link: Used to obtain percentages for Arab, Black, East Asian/Pacific Islander (adding Chinese, Korean, and Japanese percentages), Latin American, Mixed (using the percentage for "multiple visible minority"), Other (obtained by adding the Southeast Asian, Filipino, West Asian, and Minority not identified elsewhere percentages), South Asian. *Accessed on January 05, 2022*
- Census Profile for Ontario link: Used to obtain percentage of Aboriginal population. *Accessed on January 05, 2022*

- Wikipedia entry for Ontario demographics link: To corroborate that the percentage of population reported as "European" in this website matched the percentage obtained for "White Caucasian" that was independently obtained by obtaining the difference in population proportion after subtracting the sum of Visible Minorities and Aboriginal Population percentages. The aggregated information can be found in in Table A-2, where the totals per race/ethnic category are presented.

Table A-2: Reference Data from the 2016 Census from Race/Ethnicity in Ontario

| Ethnicity/Race | Percentage | Population totals |
|---|---|---|
| Arab | 1.6% | 212782 |
| Black | 4.7% | 638346 |
| East Asian/Pacific Islander | 6.6% | 886592 |
| Indigenous | 2.8% | 376558 |
| Latin American | 1.5% | 197020 |
| Mixed | 1.0% | 130033 |
| Other | 5.2% | 705333 |
| South Asian | 8.7% | 1166361 |
| White Caucasian | 67.8% | 9118079 |
| **Total** | 99.9% | 13431105 |

Visible Minorities: 29.3% Not a Visible Minority: 70.7% (Aboriginal 2.8%, White Caucasian 67.8%)

It can be seen that there are differences in the distribution of each ethnicity between the Census data and the survey. For example, Arabs/Middle Easterner correspond to 5.9% of the survey responses, but population-wise individuals that identify in this ethnic group correspond to 1.6% of the Ontario population.

### Age Groups

According to the Census, the distribution of the different the age groups for the province of Ontario are as follows:

Table A-3: Age distributions from the 2016 Census for Ontario

| Group age | Percentage | Population Totals |
|---|---|---|
| 15-24 | 12.7% | 1707959 |
| 25-34 | 12.9% | 1734856 |
| 35-44 | 12.8% | 1721407 |
| 45-54 | 14.9% | 2003826 |
| 55-64 | 13.7% | 1842444 |
| 65 and over | 16.7% | 2245898 |

To compare the survey data to the Census, the next chunk creates a barplot of the age groups in the survey. Here, it can be noticed that the age-group distribution from the survey data is different from the Census data.

### Income

Survey respondents answered the question "What is your household annual income?". To compare the distribution of responses to this question from the survey and the Census data, we used the "Household total income groups in 2015 for private households" from the Census data (available in the Census Data for Ontario website link).

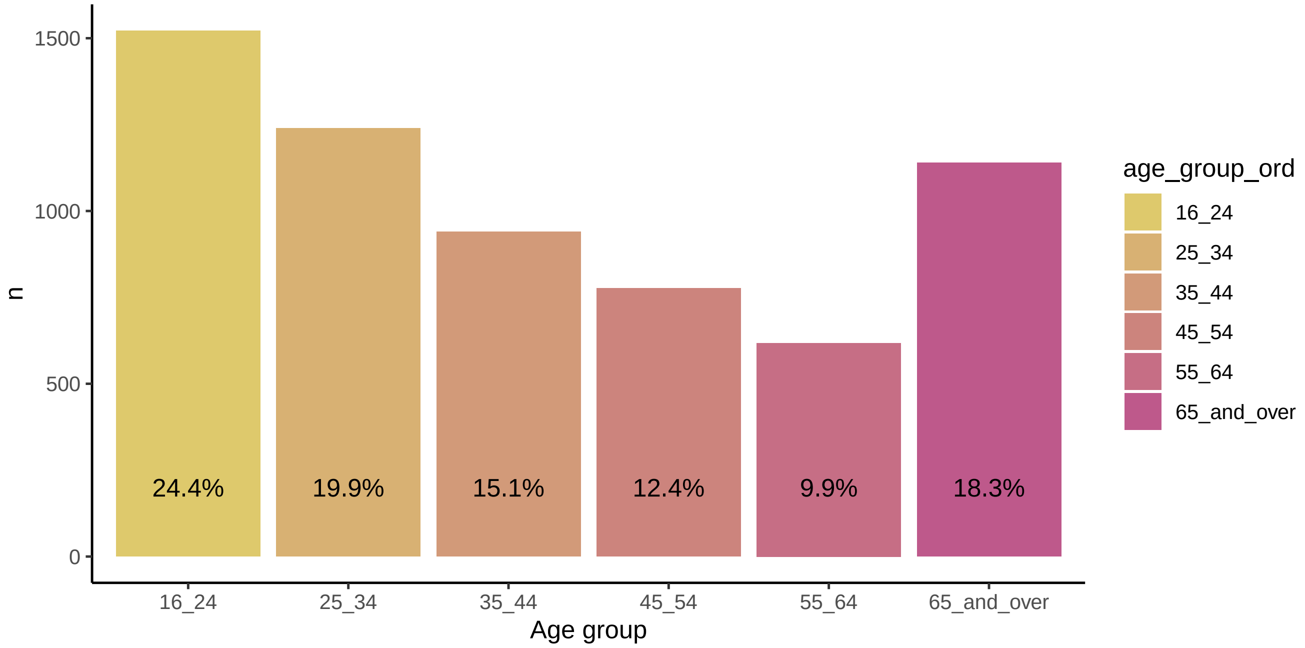


Figure A-1: Age group distributions from the dataset

Table A-4: Income percentages from the 2016 Census for Ontario

| Household income range (CAD) | Percentage | Population Totals |
|---|---|---|
| < 15,000 | 5.7% | 294643 |
| 15,000 - 24,999 | 7.5% | 387688 |
| 25,000 - 39,999 | 11.6% | 599624 |
| 40,000- 59,999 | 15.4% | 796052 |
| 60,000 - 89,999 | 19.5% | 1007988 |
| >90,000 | 40.3% | 2083176 |

One difference that is noticeable is that the brackets for income in the census data are different than the brackets used in the survey. The census does CAD 4,999 brackets (e.g., CAD 5,000- CAD 9,9999) up to CAD 49,999, followed by CAD 9,999 brackets up to CAD 99,999. After that, the brackets increase to CAD 24,999, and therefore, it is not possible to obtain percentages for the 90,000-109,999 and >110,000 brackets from the survey.

Therefore, in the following code chunk, we created an additional category for income in the dataset that matched the information from the Census, and barplots to visualize the proportion of each income bracket in the dataset.

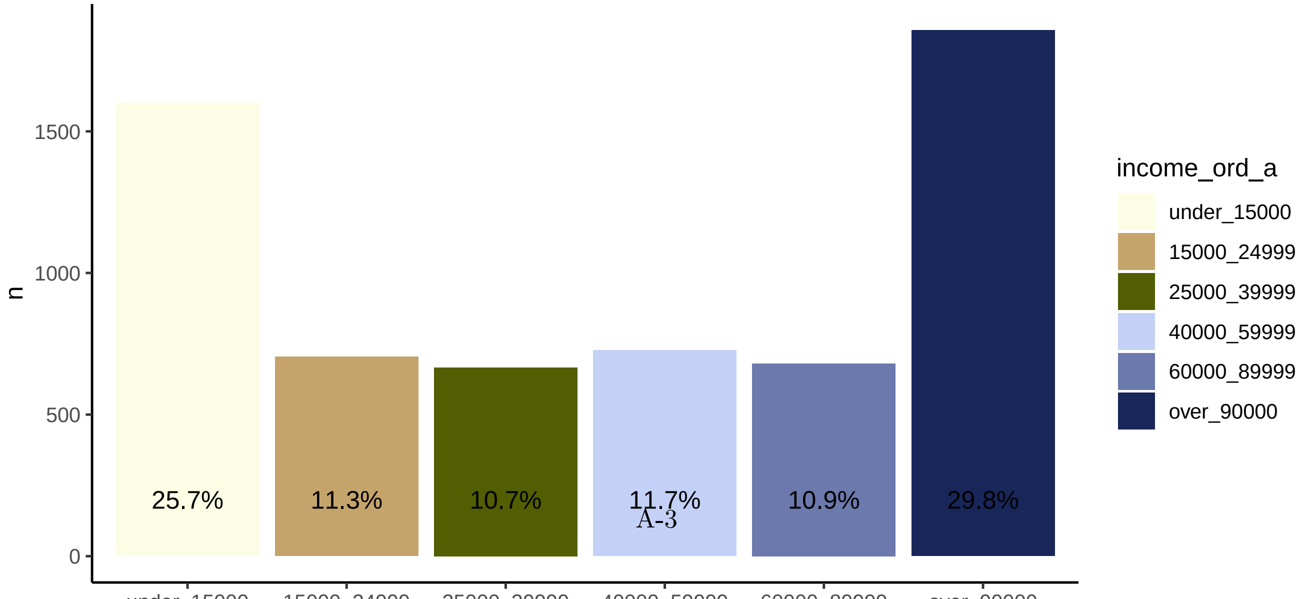

Table A-5: Population Totals for the Health Regions in Ontario

| Health Region | Population Totals |
|---|---|
| North East | 232299 |
| North West | 557000 |
| West | 4095589 |
| East | 3742520 |
| Central | 5032410 |
| Toronto | 1440644 |

Note that as indicated in the paper, we excluded observations from the North East and North West Health Regions, and therefore, the population totals for these areas were not used in the final analysis.

#### Corrections

We used the iterative proportional fitting procedure (*raking*) in order to account for the differences in income, age groups, race/ethnicity and the population from the Health Regions. Details of the implementation can be found in the R Script `raking.R` in the `code` directory.

#### Complete Model Estimates

The figures below present the complete estimates from the regression models.

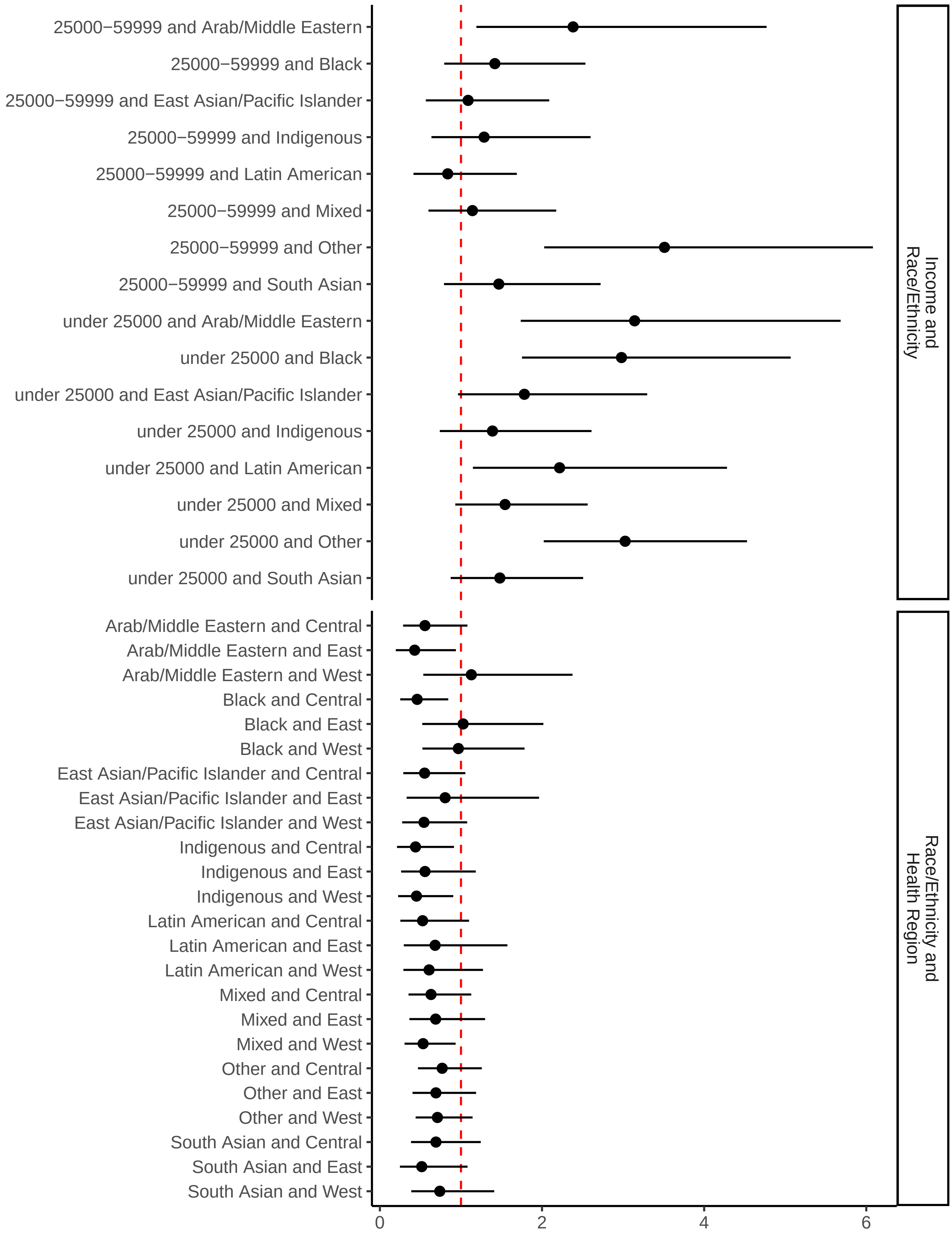


Figure A-3: Full coefficient estimates and confidence intervals for the uncorrected model.

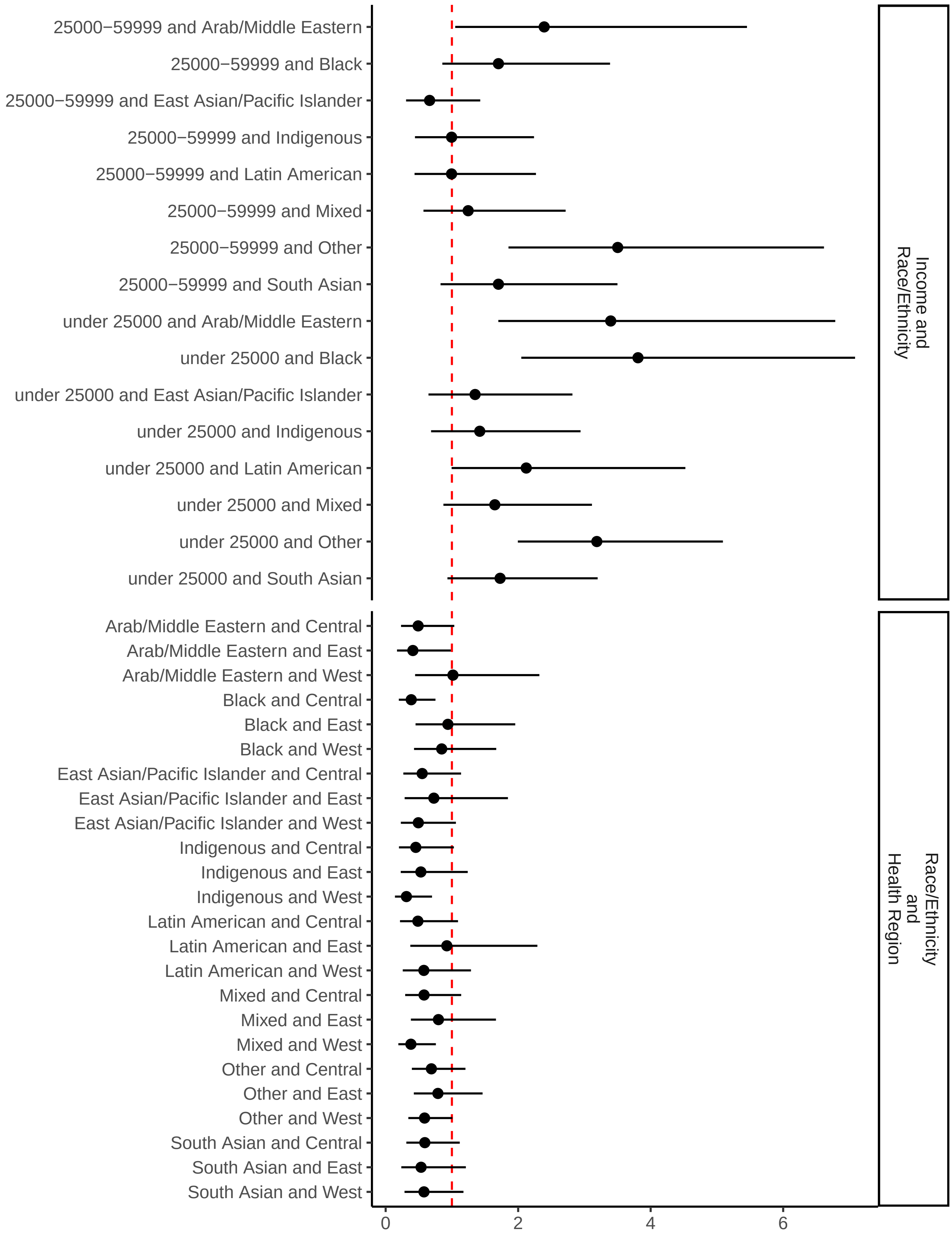


Figure A-4: Full coefficient estimates and confidence intervals for the corrected model.

#### Interactions from the Regression Model

The plots below show the interactions for Race and Income and Race and Health Region based on the corrected regression model presented in the main paper.

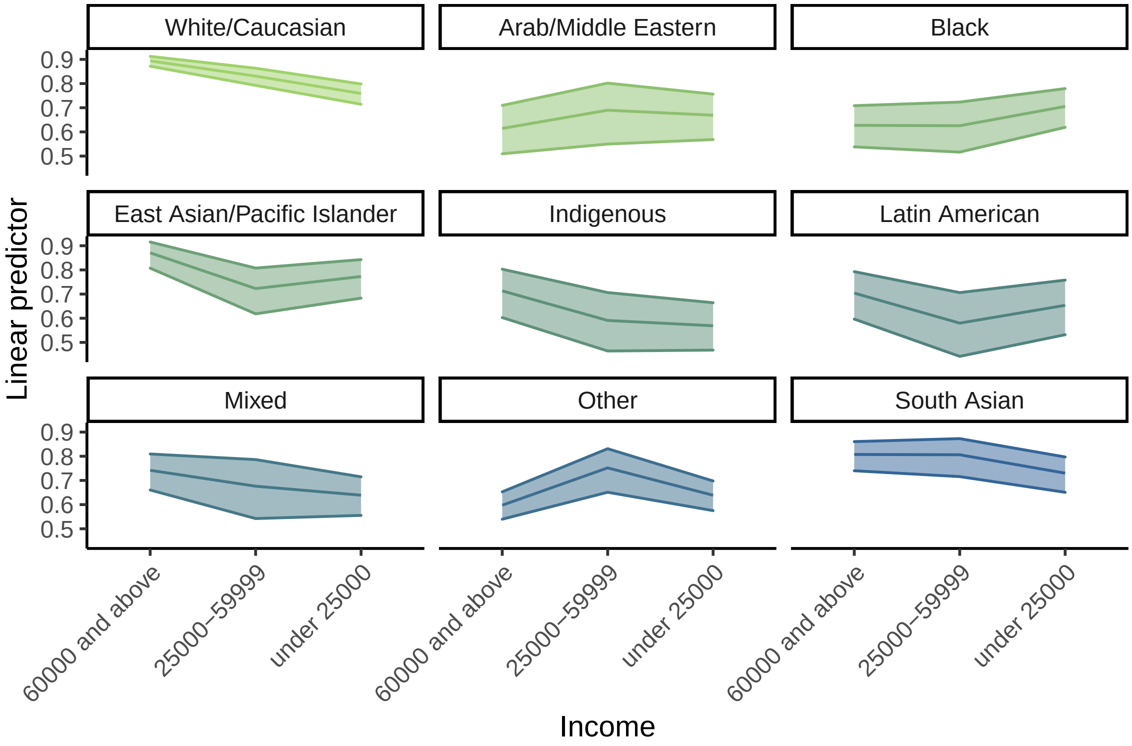


Figure A-5: Estimated marginal means for the interaction between Race and Income in the regression model.

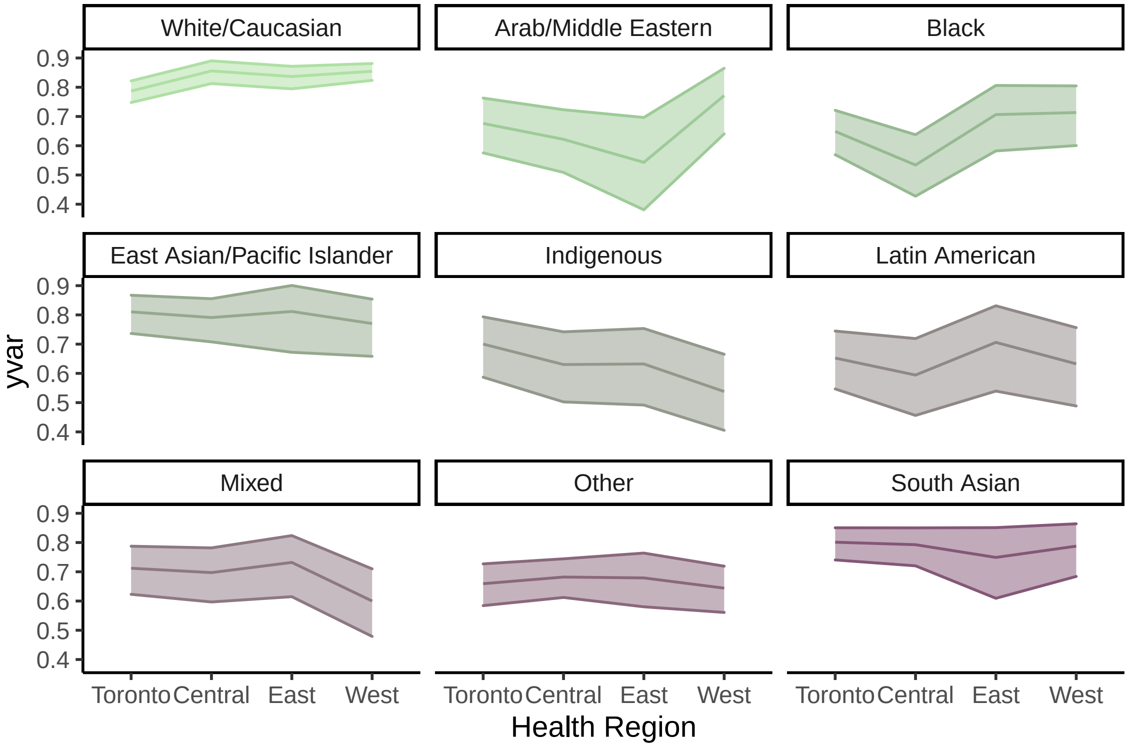


Figure A-6: Estimated marginal means for the interaction between Race and Health Region in the regression model.
